# Effects of medical school on mental health and sleep habits

**DOI:** 10.1101/2022.01.13.22269102

**Authors:** Blake McKinley, Bryan Daines, Mitchell Allen, Kayd Pulsipher, Isain Zapata, Benjamin Wilde

**Affiliations:** Department of Primary Care, Rocky Vista University College of Osteopathic Medicine, Ivins, Utah, 84738; Department of Biomedical Sciences, Rocky Vista University College of Osteopathic Medicine Parker, CO 80134

**Keywords:** Medical school, GAD-7, PHQ-9, PSQI, exercise, chronic disease, sleep quality, COVID-19

## Abstract

**BACKGROUND and OBJECTIVES:** This study aims to define changes in anxiety and depression among medical students while evaluating the association of sleep habits and other risk factors, including exercise habits and a diagnosis of chronic disease. The effect of the COVID-19 pandemic was also evaluated.

**DESIGN:** A cohort of first- and second-year medical students was evaluated longitudinally using survey methods to quantify changes from pre-medical school and summer break to each semester in medical school throughout years one and two.

**METHODS:** Data was analyzed using Generalized Linear Mixed Models (GLMMs) on the numeric responses of General Anxiety Disorder-7 (GAD-7), Patient Health Questionnaire-9 (PHQ-9), and Pittsburg Sleep Quality Index. Additional assessments evaluated exercise habits, chronic disease, and impact of COVID-19 Pandemic.

**RESULTS:** Depression, anxiety, and sleep habits displayed a cyclical change that was associated with the academic cycle. The COVID-19 pandemic was never significant. Medical students who had a chronic disease diagnosis had increased severity. Exercise did not play a role.

**CONCLUSION:** The main driver for depression, anxiety, and poor sleep quality was the academic cycle, while the COVID-19 pandemic did not have an impact on mental health.

## BACKGROUND

During the first two years of medical school, students are expected to reprioritize their lives to meet the long hours of lecture material, daily hands-on laboratory sessions, and personal study in preparation for both school and national exams [1]. To be a competitive applicant for residency, students also devote considerable time to extracurricular activities such as volunteer, work, and research experiences [2,3]. The schedule of students prior to medical school is often radically different and less demanding than that of a medical student [4,5]. These increased demands may account for the higher rates of burnout and mental illness reported in enrolled medical students compared to the general population [6–8]. Multiple studies have discovered a correlation between medical school enrollment and changes in mental health, including increased levels of depression [9–12], anxiety [6,9,12], stress [11–13], and burnout [14]. The feelings of burnout and depression that present in medical students also persist in medical residents and physicians [10,15]. Therefore, the ability to identify risk factors that contribute to mental health deterioration in medical students is likely to have implications for residents and physicians.

Risk factors which impact mental health include sleep quality and quantity, exercise, and chronic disease [16–18]. Arguably, poor sleep quality has the most significant association with poor mental health in medical students. It has been shown to be associated with an increase in stress, anxiety, and depression among medical students [12,13,19]. College students that present with depressive symptoms were found to have greater comorbid anxiety when they also experienced sleep disturbances [12,19–21]. Students with depressive symptoms were found to rate sleep quality as poor and experience increased somnolence in class [22]. Poor sleep quality, decreased quantity, and increased depression levels were shown to be independent predictors of burnout [14] and have a strong association with decreased mental health. Additionally, reduced incidence rates of depression and some anxiety disorders have been observed in exercising individuals [23]; exercise has even been shown to be effective in the treatment of depression, and to a lesser extent anxiety [23,24]. Likewise, chronic disease has been shown to lead to depression and anxiety [17]. Adding to the equation, the COVID-19 pandemic came into the picture. Depression, anxiety, and sleep disorders have been linked to the pandemic among the general public [25]. More specifically, studies have noted that college students are anxious regarding cancellation of examinations and academic events [26]. The effects of these disruptions are likely to contribute negatively to students’ mental health.

Anxiety and depression were chosen as metrics to evaluate mental health along with sleep quality due to its strong association with them. The purpose of this study was to define changes in anxiety, depression, and sleep quality of medical students in their first two years of medical school while identifying associated risk factors. The following risk factors were considered: chronic disease, sleep quantity, year of medical school, class year, exercise habits, and COVID-19 pandemic. This allows medical schools to better understand the effect that medical school has on their students, enabling them to develop preventative and coping strategies to help students minimize mental distress. Additionally, by determining the effectiveness of assessment tools and their association to risk factors, it will be possible to have reliable methods to monitor and to detect students at risk. Medical students will become future physicians that will provide an important service to our society. It is important to improve the well-being of medical students so they can become successful physicians.

## METHODS

### Participants and study approach

An invitation to participate in this study was sent out to all enrolled students in the class of 2023 and the following year to the class of 2024 in a medical school with two separate campuses. Follow-up survey invitations were emailed to those students that signed the informed consent to participate in this study and completed the first survey. An outline of the study is presented in Figure 1.

**Figure 1.**
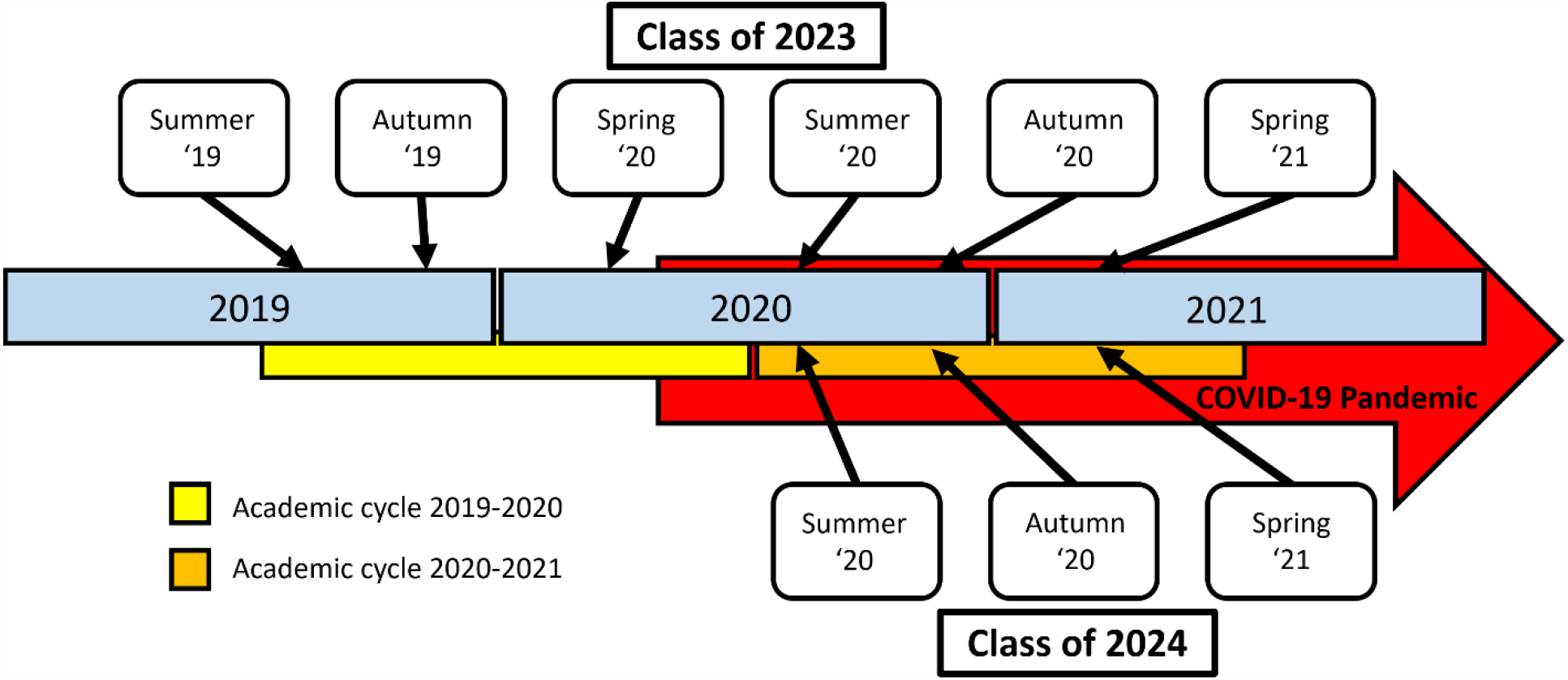
Study timeline.

No compensation was provided for participating in the study and all participation was voluntary. All participants signed an electronic consent form prior to participating. The research project was reviewed by the Rocky Vista University Institutional Review Board with the corresponding IRB# 2019-0070. The research project was deemed exempt from a full IRB review. No specific funding was received to conduct this study.

### Surveys

Surveys were designed to assess underlying chronic disease, sleep quantity and quality, exercise habits and levels of depression and anxiety. A chronic disease was specified as one diagnosed by a physician. Sleep quantity was assessed as average hours of sleep per night. Sleep quality was assessed the first year of the study with three questions that incorporated graded responses to quantify sleep quality changes (SQ-3). Although the sleep quality questions were common questions used in other studies to quantify sleep quality, they have never been used in this format. The Pittsburgh Sleep Quality Index (PSQI) [27] was added starting with the survey for the Summer of 2020 to supplement the evaluation of sleep quality. The original three questions assessing sleep quality remained as a part of the survey to be computed separately from PSQI. Exercise was assessed as days per week and minutes per session. The total minutes of exercise per week was calculated by multiplying these two values. Finally, to assess mental health, validated questionnaires were used to analyze levels of anxiety and depression, Generalized Anxiety Disorder-7 (GAD-7) and Patient Health Questionnaire-9 (PHQ-9), respectively [28,29]. Therefore, all subjective measurements: sleep quality, anxiety levels, and depression levels were assessed with previously validated questionnaires. Additional questions were also added in year two of the study to evaluate for self-reported effects of COVID-19, starting with the summer survey of 2020. Specifically, questions were added to assess if students believed that COVID-19 had increased their levels of anxiety, depression, and/or decreased quality of sleep. An example of the survey used is available as a supplementary file.

### Statistical Analysis

Data was analyzed using Generalized Linear Mixed Models (GLMMs) on the numeric responses of SQ-3, GAD-7, PHQ-9 and PSQI. These models included fixed effects of class year (class of 2023/class of 2024), COVID-19 period (Pre-/ Post-), specific time points (Summer ‘19 through Spring ‘21), chronic disease diagnosis (Yes/No), average number of sleep hours (hours), and total exercise per week (minutes). Repeated measurements by participants were included in the models as a random effect. All models assumed errors to be normally distributed with mean zero and variance of σ^2^. All modeling and descriptive statistical evaluations were performed with SAS v.9.4 (SAS Institute, Cary, NC). Statistical significance was declared at a confidence level of 95%.

The analysis was performed on three separate cohorts defined by the time points and class years. The first modeling analysis was performed on the full dataset which included both classes of 2023 and 2024, this cohort did not analyze COVID-19 specific questions. The second modeling analysis did include the COVID-19 specific questions. The third, and last modeling analysis, was performed only on the class of 2023 cohort, this was done to separate the effect of participants from the class of 2024 that had only been in medical school during the pandemic.

## RESULTS

The total number of participants in the study was 197; where 85 out of 130 students participated from the class of 2023 and 112 out of 145 students participated from the class of 2024. Participant frequencies are summarized in Table 1 by COVID period, time point, and chronic disease diagnosis. A summary for the mean and standard deviation of all variables evaluated in this study is presented in Table 2. In this table, all values are presented for the full dataset, per class year cohort, per COVID-19 period, and by chronic disease diagnosis. Among all participants 27.73% had at least one missing record for one or more timepoint.

**Table 1.**
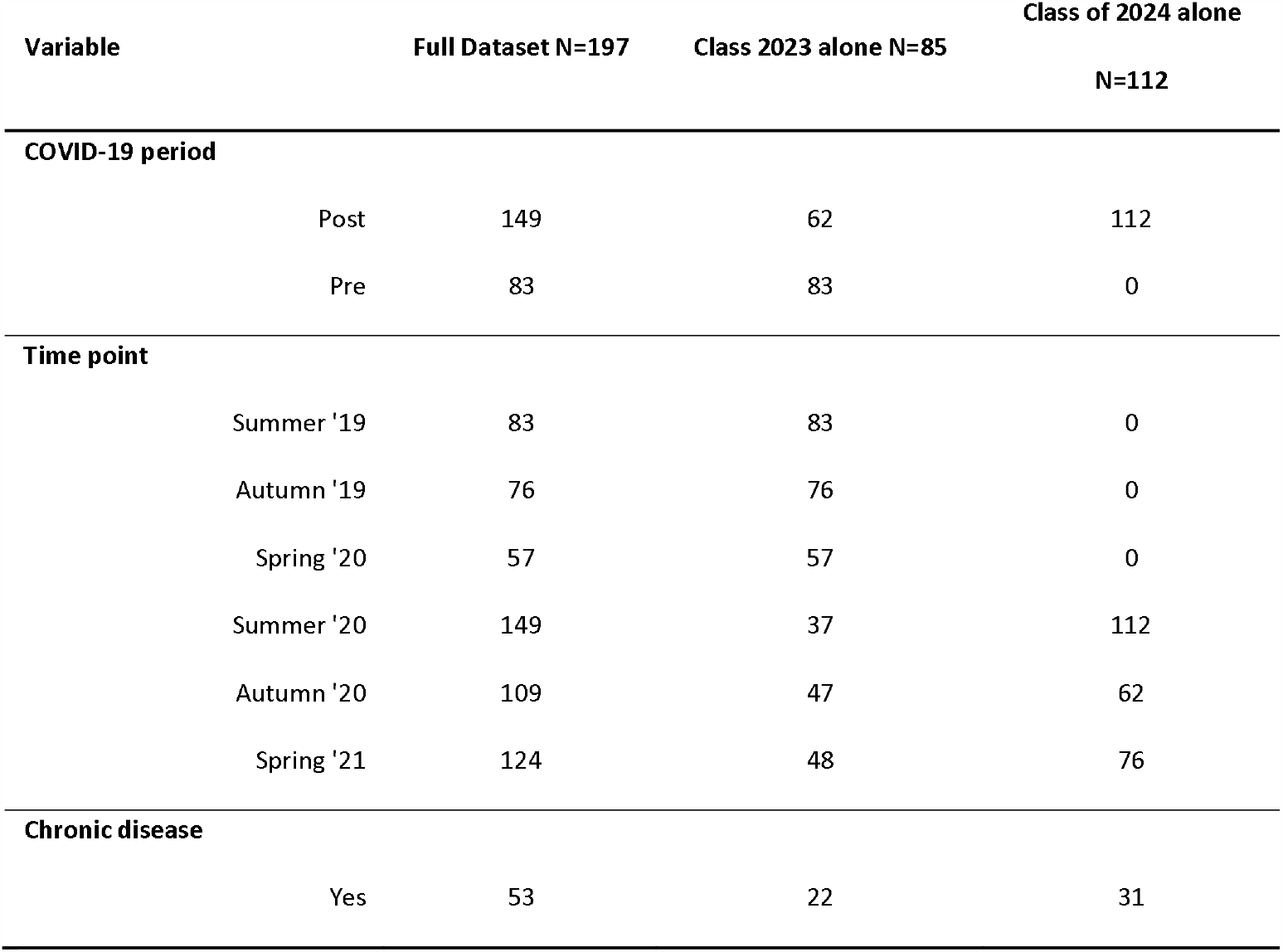
Participants included in the study on the full dataset and by class year cohorts.

**Table 2.** Mean and Standard Deviations in the dataset.

| Variable | Full dataset |  | Class year |  |  |  | COVID-19 Period |  |  |  | Chronic disease |  |  |  |
| --- | --- | --- | --- | --- | --- | --- | --- | --- | --- | --- | --- | --- | --- | --- |
|  |  |  | 2023 |  | 2024 |  | Pre |  | Post |  | Yes |  | No |  |
|  | Mean | Std Dev | Mean | Std Dev | Mean | Std Dev | Mean | Std Dev | Mean | Std Dev | Mean | Std Dev | Mean | Std Dev |
| Average Sleep Hours<br>(Hours) | 7.474 | 1.129 | 7.799 | 1.059 | 7.024 | 1.068 | 7.799 | 1.065 | 7.043 | 1.067 | 7.425 | 1.213 | 7.496 | 1.089 |
| SQ-3 | 5.830 | 3.003 | 5.776 | 3.060 | 5.908 | 2.923 | 5.769 | 3.083 | 5.914 | 2.893 | 6.758 | 2.707 | 5.539 | 2.962 |
| Total exercise per week<br>(Minutes) | 201.17 | 177.33 | 190.91 | 162.47 | 215.52 | 195.69 | 190.91 | 163.59 | 214.92 | 193.72 | 198.15 | 203.65 | 202.57 | 163.98 |
| GAD-7 | 6.035 | 5.057 | 5.701 | 5.036 | 6.508 | 5.060 | 5.719 | 5.024 | 6.465 | 5.081 | 7.651 | 5.609 | 5.408 | 4.608 |
| PHQ-9 | 5.127 | 5.055 | 5.065 | 5.084 | 5.216 | 5.022 | 5.029 | 5.013 | 5.262 | 5.119 | 7.016 | 5.878 | 4.362 | 4.399 |
| COVID-19 Depression | -0.128 | 1.291 | -0.150 | 1.264 | -0.116 | 1.307 | -0.157 | 1.263 | -0.113 | 1.307 | 0.252 | 1.272 | -0.316 | 1.261 |
| COVID-19 Anxiety | 0.232 | 1.366 | 0.128 | 1.305 | 0.288 | 1.396 | 0.157 | 1.318 | 0.270 | 1.390 | 0.583 | 1.288 | 0.059 | 1.372 |
| COVID-19 Quality of<br>sleep | -0.668 | 1.124 | -0.702 | 1.155 | -0.648 | 1.109 | -0.672 | 1.162 | -0.665 | 1.106 | -0.447 | 1.095 | -0.780 | 1.124 |
| PSQI | 5.864 | 3.190 | N/A | N/A | 5.940 | 3.195 | N/A | N/A | 5.902 | 3.181 | 7.291 | 3.185 | 5.150 | 2.949 |

The GLMM modeling analysis used in this study allows for simultaneous evaluation of effects such as class year, COVID-19 period, differences across time points, chronic disease, average sleep hours, and total exercise. Therefore, estimation for individual effects on SQ-3, GAD-7, PHQ-9, and PSQI is already adjusted for other effects in the model. Figure 2 presents the differences across time points for the study’s full dataset. Significant differences were observed across time points in the individual analysis of SQ3, GAD-7, PHQ-9, and PSQI. When examining the differences across time point results for SQ-3, GAD-7, PHQ-9, PSQI, it is evident that there are similar trends. Pre-medical school for class of 2023 (Summer ‘19); and pre-medical school for class of 2024, combined with class of 2023 who were on summer break before starting year 2 of medical school (Spring ‘21), reported significant lower scores for all time points. The differences across time points, a lower number of sleep hours and having a diagnosis of chronic diseases significantly increased SQ-3, PHQ-9 and PSQI. The differences across time points and having a diagnosis of chronic diseases significantly increased GAD-7. P-values for these associations are presented in Table 3. A separate evaluation of timepoints (pre-medical school, fall semester, and spring semester of year one) for individual class year cohorts (class of 2023 and class of 2024) displayed the same pattern (data not shown).

**Figure 2.**
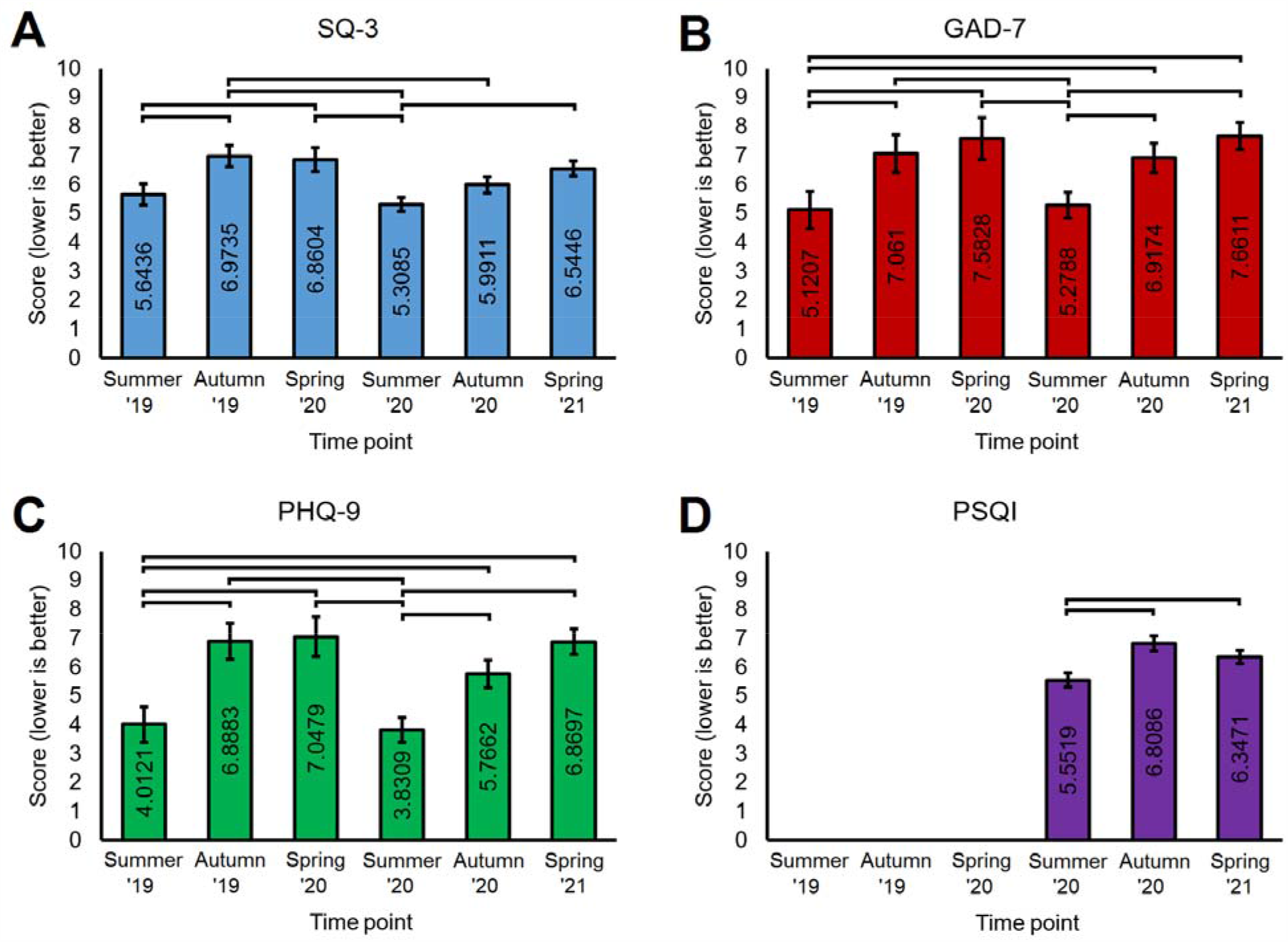
Time point effect estimates. A) SQ-3. B) GAD-7. C) PHQ-9. D) PSQI. In all panels, brackets indicate pairwise differences at a P≤0.05 level. Error bars correspond to the Standard Error of Mean (SEM).

**Table 3.**
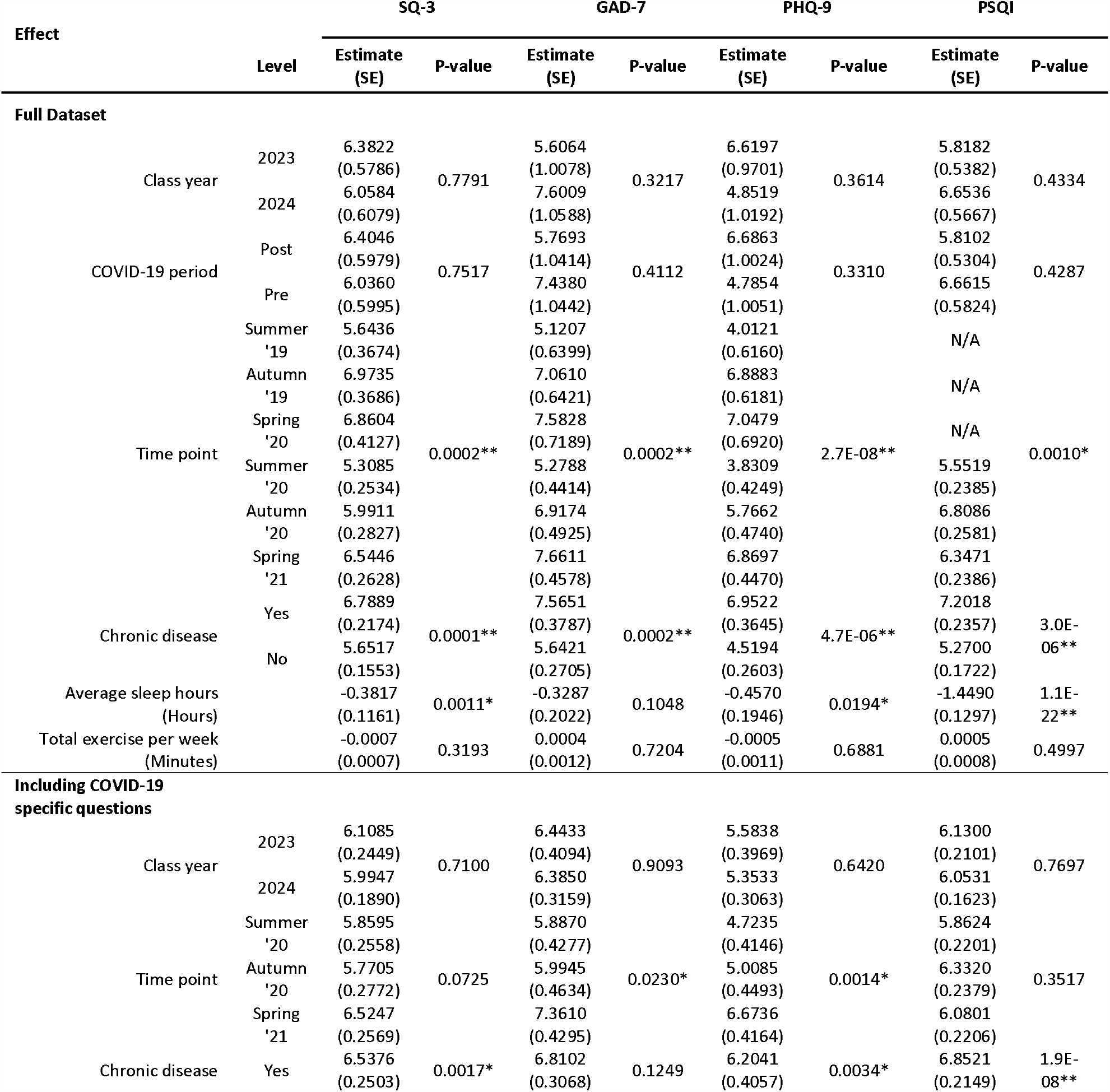
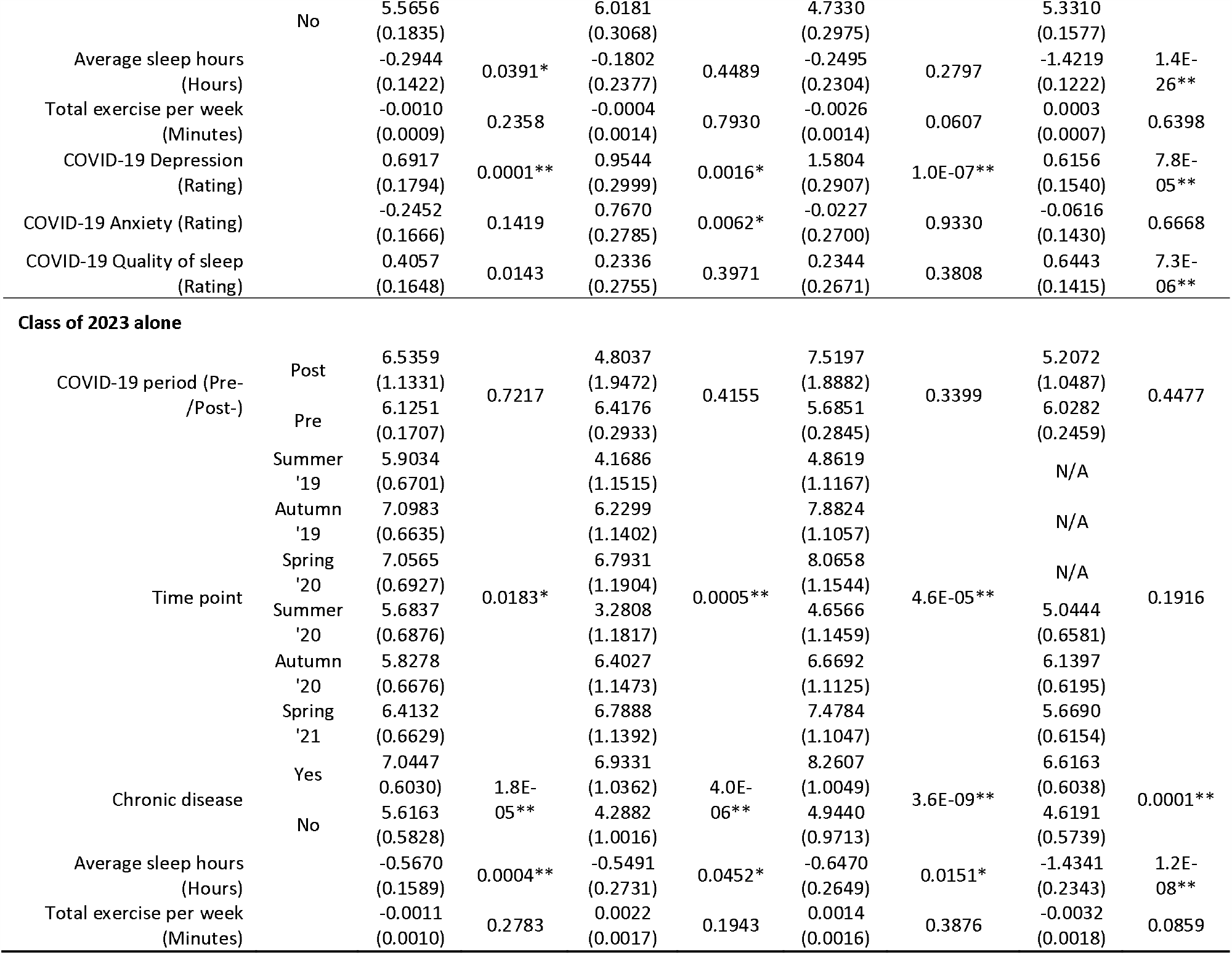
Estimates and significance testing for each model’s included effects. Significant associations are declared at P≤0.05 and are indicated in the table with an asterisk *. Bonferroni corrected significant associations are declared at P≤6.5E-4 and are indicated with a double asterisk **. Columns correspond to the assessment tool evaluated while rows correspond to the p-value of such association. Outcomes for the three models are presented: full dataset, dataset including COVID-19 specific questions and class of 2023 alone.

The full data was then run again in a similar fashion but included the three additional variables of self-reported change of anxiety, depression, and sleep quality due to COVID-19; labeled as “COVID-19 Anxiety,” “COVID-19 Depression,” and “COVID-19 Quality of sleep”. With this addition, the self-reported COVID-19 variables provided more information by displaying significant associations to SQ-3, GAD-7, PHQ-9 and PSQI scores; differences across time points had a significant effect on GAD-7 and PSQI; having a chronic disease diagnosis had a significant effect on SQ3, PHQ-9, and PSQI; average sleep hours had a significant effect on SQ-3 and PSQI; COVID-19 Depression had a significant effect on SQ-3, GAD-7, PHQ-9, and PSQI; COVID-19 Anxiety had a significant effect on GAD-7; and COVID-19 Quality of sleep had a significant effect on SQ-3 and PSQI. No other variables had a significant effect. P-values for these associations are presented in Table 3.

Last, the data was analyzed again in a similar manner but only for the class of 2023, excluding all data from the class of 2024, with variables of COVID-19 period, differences across time points, chronic disease, average sleep hours, and total exercise accounted for when assessing for the individual effect on SQ3, GAD-7, PHQ-9, and PSQI. Time point change had a significant effect on SQ-3, GAD-7, PHQ-9; having a chronic disease diagnosis and average sleep hours had a significant effect on SQ-3, GAD-7, PHQ-9, and PSQI. No other variables had a significant effect. P-values for these associations are presented in Table 3.

In summary, the evaluations of differences across time points, along with the consideration of the effect of the COVID-19 pandemic, that was never significant, suggests that: life as a medical student (with the accompanying stress that accumulates through the academic year), having a chronic disease diagnosis, and fewer hours of sleep are the evident causes of increased levels of anxiety, depression, and a decrease in sleep quality.

## DISCUSSION

Medical school has a significant effect on sleep quality, depression and anxiety [9–14,19,21,30–35]. The majority of analyses that have been performed are cross sectional studies that are an assessment of one time point, relying on a comparison of their results against other groups to provide context of the significance of their findings, including the general population [6,30,35], or to other assessments of medical students from around the world [5,12,30,31,36]. However, the longitudinal analysis of students in this study is unique in that it allowed the quantification of the change from pre-medical school and summer break to each semester in medical school throughout years one and two. This allowed students to act as their own controls, reducing for confounding factors. After completing the assessment, the differences across time points displayed a cyclic shift in negative mental health and poor sleep quality of medical students that was associated directly with the academic year.

There is great variability in results of sleep quality, depression, and/or anxiety of medical students. The reported percentages of students that suffer from poor sleep quality range from 55.8-76%, anxiety from 41-73%, and depression from 10.4-65% [11–13,19,21,30–33,36]. This variation is due in part to the lack of standardized assessment tools designated to assess anxiety, depression, and sleep quality in medical students, leading to a variety of assessment tools used. Additionally, the different structures and expectations within medical schools around the world are difficult to account for. For that reason, this study does not focus on comparing the percentages of total anxiety, depression, and sleep quality to other analyses, but assesses the longitudinal change.

As mentioned, school structure influences results. The medical school assessed in this study is structured similarly to the general model of medical schools throughout the United States: 4-year curriculum, divided into two-years of basic science coursework and two-years of clinical clerkships [37]. This study evaluated students in their first two years of basic science coursework, as third-and-fourth years change in structure and setting. No significant difference in anxiety, depression, and/or sleep quality was observed between year one and year two. However, differing levels of anxiety, depression, and sleep quality have been observed in other studies when all years of training are considered [32,38].

The analysis revealed that class year (class of 2023 versus class of 2024) and exercise habits (computed as minutes per week) had no effect on mental health or sleep quality. Although exercise has shown to cause reduced incidence of anxiety and depression in the general population [16], this study suggests that there is no significant effect in medical students. However, this does not rule out the effect that adding more exercise may have on students found to have anxiety and depression, as exercise has shown to have therapeutic effects on mental health [23].

The presence of chronic disease, diagnosed by a physician (self-reported), was shown to lead to higher anxiety and depression levels while decreasing sleep quality. A systematic review which included 20 studies outlined that chronic disease may be interrelated to anxiety/depression, one causing the other or both potentially exacerbating each other [17]. A simple explanation for this finding could be that a medical student only can compensate or adjust so much. Thus, when a student is living with a chronic illness, their health is already compromised; when the pressures of medical school are added, they are limited in their reserve to compensate and are therefore more susceptible to developing increased levels of anxiety and depression.

In general, fewer hours of sleep were also shown to have a strong negative effect on sleep quality while also negatively affecting mental health. Number of hours of sleep was shown to significantly affect depression levels more so than anxiety levels. Evidence suggests that sleeping less than six hours leads to the development of chronic disease [39], a risk factor for increased anxiety, depression, and poor sleep quality. One study found that medical students often sacrifice sleep to meet the demands of school [13]. Helping students prioritize sufficient hours of sleep each night could have long term benefits.

This analysis provides insight into areas that medical schools should place focus, including sleep patterns and mental health. It has been shown that “good” sleep improves academic performance [40] and that medical students who are sleep deprived have significantly lower GPAs [41]. Anxiety, depression, and stress lead to poor academic performance in medical students [42,43]. Students often can recognize these problems and perceive its effect on their academic performance [22], but do not always know what to do to receive help. At a United States medical school where confidential mental health services were available, only 26.5% of the identified depressed students reported treatment. Notably, 20.4% of the depressed students reported experiencing suicidal ideation [33].This highlights that the overall well-being of students should be considered.

Medical school should have a focus on health promotion with educational programs that emphasize the importance of sleep and mental health [21]. One suggestion to help prevent depression is to educate students on appropriate sleep hygiene and encourage students to pursue professional guidance to treat sleep disturbances [22]. Another suggestion to reduce mental health symptoms is for medical schools to institute stress coping mechanisms and improvement of interpersonal relationships [11]. There is also advice on how to establish a well-being curriculum [44].

### Assessment of the impact of COVID-19

The COVID-19 pandemic arose about halfway through the data collection period of this study and changed educational delivery dramatically during this time, incorporating a virtual learning environment to encourage social distancing [45,46]. Studies have shown that the pandemic has had a negative impact on depression, anxiety, and sleep disorders [25,26]. However, there are studies that report increases in sleep among college students, [47].

In this study the effect of COVID-19 was evaluated in two ways: first, by evaluating directly its effect in the model (pre- and post-COVID-19 period variable) and second, by comparing the first three time points (Summer ‘19 - Spring ‘20), representing student results from the summer, fall semester, and spring semester of the school year 2019-2020 (pre-COVID-19), to the corresponding seasons of the following year: time points 4-6 (Summer ‘20 - Spring ‘21), representing student results from the summer, fall semester, and spring semester of the school year 2020-2021 (post-COVID-19). On the full dataset, there were no differences across time points for the same season of the year and there was no effect on the change from pre- to post-COVID-19 period variable. This suggests that there was no significant contribution from the COVID-19 pandemic to anxiety, depression, or sleep quality. To further account for possible unaccounted confounding effects of putting together the two classes (2023 and 2024 together) the analysis was also run with only the class of 2023. Findings also revealed that the COVID-19 period had no significant effect on any of the scores. These findings agree in part with a recent report on college students where the effect of the COVID-19 pandemic on stress and sleep was not necessarily negative and suggested that students are largely resilient [48]. After COVID-19 pandemic started, 3 questions were added to the surveys to account for any self-reported effect of COVID-19 on sleep quality, anxiety, and depression. The results were significant for worse outcomes in all three categories. Change in anxiety, depression, and sleep quality as assessed by GAD-7, PHQ-9, and SQ-3, respectively, were not significant, but the self-reported responses were. These findings suggest that students that are unable to recognize the cause of their decline in mental health are likely to blame the pandemic more than the contribution of their life as a medical student.

### Limitations and future directions

A limitation of this study can be its representativeness since data was collected from a single medical school consisting of two campuses. Also, the sleep quality analysis added an additional assessment, PSQI, that was not available for the first year. Future research should be directed at expanding the medical student population surveyed across multiple universities to better generalize results. Furthermore, studies should incorporate medical students in years three and four to provide data that spans the entire medical school training.

## CONCLUSION

The first two years of medical school is associated with elevated levels of depression, anxiety, and poor sleep quality; the academic cycle being the most important driver. Chronic medical conditions and decreased total number of sleep hours were identified as risk factors for poor mental health and poor sleep quality among medical students. The COVID-19 pandemic did not have an impact on medical student mental health or sleep patterns. Medical schools and students themselves should take an active role in addressing sleep patterns and mental health to achieve overall better well-being, improved academic performance, and ultimately become better physicians.

## Supporting information

Supplementary file Survey example

## Data Availability

All data produced in the present study are available upon reasonable request to the authors

