## Supplementary file Survey example for "Effects of medical school on mental health and sleep habits"

**Thank you for participating in our survey. Your feedback is important.**

**This survey contains questions regarding sensitive mental health topics such as depression, anxiety and burnout. If you or someone you know is struggling with any of these issues, the following resources are available for you:**

**Kathy Killian-Harmon, LMFT**
**Mental Health and Wellness Counselor**
**Rocky Vista University, Ivins, UT**
**Cell: 435-668-7996**

**Student Linc: Available 24/7 every day of the year. Every call is answered by licensed clinical professional.**

**24 Hour Suicide Prevention and Mental Health Crisis Lifeline**

1. **Please read the consent form below. Sign the consent form by typing your name in the box below. Thank you.**

RESEARCH PARTICIPANT Consent Form

Health Patterns of Medical Students Before and After Matriculation to Medical School.

You are invited to be in a research study of seeking to understand the influence of sleep and exercise patterns on physical, mental health, and academic success. We ask that you read this form and ask any questions you may have before agreeing to be in the study. Your participation is entirely voluntary.

This study is being conducted by Bryan Daines, Blake Mckinley, Kayd Pulsipher, and Mitchell Allen, students at Rocky Vista University. Under the direction of Ben Wilde, Department Vice Chair of Clinical Medicine, Rocky Vista University.

Purpose of Study

The purpose of this study is to understand the influence of sleep and exercise patterns on physical and mental health and performance of medical students at RVU, comparing pre-medical school to that of medical school. You were selected as a prospective participant in this study because you are a student at Rocky Vista University.

Procedures

If you agree to be in this study, we will ask you to do the following things:

You as a participant will be asked to truthfully complete multiple surveys throughout the year. You as a participant will be individually identified via email to track individual progression.

Duration

Participation in the study involves the following time commitment:

Completion of each survey will take an estimated time of 5-10 minutes. You as a participant will be asked to complete a survey in September 2020, in January 2021, and June 2021.

Risks and Benefits of Being in the Study

The study involves the following foreseeable risks and/or discomforts:

There is very low risk for you as a participant in this study. We will be identifying participants by obtaining an email address. This will allow us to send follow up surveys to track individual change in students. We will also be asking about medical conditions and class rank. However, these results will be kept confidential, and Dr. Wilde will be the only researcher with the email associated with your answer responses.

The benefits to participation are:

The results of this study could better help students identify their current health patterns; and possibly correct these patterns to positively impact future academic performance, and long-term mental and physical health. These results could help identify risk factors for medical students before matriculation medical school.

Compensation and/or Costs

There will be no compensation for your participation in this study, but your help is appreciated by the researchers.

Confidentiality

Participation in research may involve some loss of privacy; however, the researchers will make every effort to ensure that information about you remains confidential. Your identity will not be revealed in any publications, presentations, or reports resulting from this research study. However, it may be possible for someone to recognize your particular story/situation/response. Student researchers will be unable to have access to the email responses by the participates, therefore eliminating the possibility of your specific name associated to your responses.

We will collect your information through a survey via Survey Monkey, which is an online survey development cloud-based software company. This information/data will be stored and safeguarded by a password protected, cloud-based storage system.

**Your identifiable private information collected as part of the research will not be used or distributed for future studies.**

Please print your name as consent to participate in this survey:

2. What is your email address?

3. Do you have a medically diagnosed chronic disease, disorder, or illness?

- Yes
- No

4. What is the condition?

5. How often do you experience symptoms from your condition?

- Never
- <1 time per week
- 1-6 times per week
- 1-2 times per day
- >2 times per day
- Constantly
- N/A

6. Do you feel like this medical condition negatively affects your overall energy levels?

- Yes
- No
- N/A

7. How many years ago were you diagnosed with this condition?

0 — 50 [ ]

**How often are you currently bothered by the following problems?**

8. Feeling nervous, anxious or on edge

- 0- Not at all
- 1 - Several Days
- 2- More than half the days
- 3- Nearly every day

9. Not being able to stop or control worrying

- 0 - Not at all
- 1 - Several Days
- 2 - More than half the days
- 3 - Nearly every day

10. Worrying too much about different things

- 0 - Not at all
- 1 - Several Days
- 2 - More than half the days
- 3 - Nearly every day

11. Trouble relaxing

- 0 - Not at all
- 1 - Several Days
- 2 - More than half the days
- 3 - Nearly every day

12. Being so restless that it's hard to sit still

- 0 - Not at all
- 1 - Several Days
- 2 - More than half the days
- 3 - Nearly every day

13. Becoming easily annoyed or irritable

- 0 - Not at all
- 1 - Several Days
- 2 - More than half the days
- 3 - Nearly every day

14. Feeling afraid as if something awful might happen

- 0 - Not at all
- 1 - Several Days
- 2 - More than half the days
- 3 - Nearly every day

15. Little interest or pleasure in doing things

- 0 - Not at all
- 1 - Several Days
- 2 - More than half the days
- 3 - Nearly every day

16. Feeling down, depressed, or hopeless

- 0 - Not at all
- 1 - Several Days
- 2 - More than half the days
- 3 - Nearly every day

17. Trouble falling or staying asleep, or sleeping too much

- 0 - Not at all
- 1 - Several Days
- 2 - More than half the days
- 3 - Nearly every day

18. Feeling tired or having little energy

- 0 - Not at all
- 1 - Several Days
- 2 - More than half the days
- 3 - Nearly every day

19. Poor appetite or overeating

- 0 - Not at all
- 1 - Several Days
- 2 - More than half the days
- 3 - Nearly every day

20. Feeling bad about yourself or that you are a failure or have let yourself or your family down

- 0 - Not at all
- 1 - Several Days
- 2 - More than half the days
- 3 - Nearly every day

21. Trouble concentrating on things, such as reading the newspaper or watching television

- 0 - Not at all
- 1 - Several Days
- 2 - More than half the days
- 3 - Nearly every day

22. Moving or speaking so slowly that other people could have noticed. Or the opposite, being so figety or restless that you have been moving around a lot more than usual

- 0 - Not at all
- 1 - Several Days
- 2 - More than half the days
- 3 - Nearly every day

23. Thoughts that you would be better off dead, or of hurting yourself

- 0 - Not at all
- 1 - Several Days
- 2 - More than half the days
- 3 - Nearly every day

24. Have you met with a mental health group or psychologist to help with anxiety or depression?

- Yes
- No

25. Do you take any medications for anxiety or depression?

- Yes
- No

26. COVID-19 has caused me to experience higher levels of depression currently.

- Strongly agree
- Agree
- Neither agree nor disagree
- Disagree
- Strongly disagree

27. COVID-19 has caused me to experience higher levels of anxiety currently.

- Strongly agree
- Agree
- Neither agree nor disagree
- Disagree
- Strongly disagree

28. During the past month, what time have you usually gone to bed at night?

Bed time:

Time AM/PM

hh mm -

29. During the past month, how long (in minutes) has it usually taken you to fall asleep each night?

Number of minutes:

30. During the past month, what time have you usually gotten up in the morning?

Getting up time:

Time AM/PM

hh mm -

31. During the past month, how many hours of actual sleep did you get at night? (This may be different than the number of hours you spent in bed.)

Hours of sleep per night:

**For questions 32-42, check the one best response. Please answer all questions.**

**During the past month, how often have you had trouble sleeping because you...**

32. Cannot get to sleep within 30 minutes

- Not during the past month
- Less than once a week
- Once or twice a week
- Three or more times a week

33. Wake up in the middle of the night or early morning

- Not during the past month
- Less than once a week
- Once or twice a week
- Three or more times a week

34. Have to get up to use the bathroom

- Not during the past month
- Less than once a week
- Once or twice a week
- Three or more times a week

35. Cannot breathe comfortably

- Not during the past month
- Less than once a week
- Once or twice a week
- Three or more times a week

36. Cough or snore loudly

- Not during the past month
- Less than once a week
- Once or twice a week
- Three or more times a week

37. Feel too cold

- Not during the past month
- Less than once a week
- Once or twice a week
- Three or more times a week

38. Feel too hot

- Not during the past month
- Less than once a week
- Once or twice a week
- Three or more times a week

39. Had bad dreams

- Not during the past month
- Less than once a week
- Once or twice a week
- Three or more times a week

40. Have pain

- Not during the past month
- Less than once a week
- Once or twice a week
- Three or more times a week

41. Other reason(s), please describe

42. If other reason listed in Question 41, then how often during the past month have you had trouble sleeping because of this?

- Not during the past month
- Less than once a week
- Once or twice a week
- Three or more times a week

43. During the past month, how would you rate your sleep quality overall?

- Very good
- Fairly good
- Fairly bad
- Very bad

44. During the past month, how often have you taken medicine to help you sleep (prescribed or "over the counter")?

- Not during the past month
- Less than once a week
- Once or twice a week
- Three or more times a week

45. During the past month, how often have you had trouble staying awake while driving, eating meals, or engaging in social activity?

- Not during the past month
- Less than once a week
- Once or twice a week
- Three or more times a week

46. During the past month, how much of a problem has it been for you to keep up enough enthusiasm to get things done?

- No problem at all
- Only a very slight problem
- Somewhat of a problem
- A very big problem

47. Do you have a bed partner or room mate?

- No bed partner or room mate
- Partner/room mate in other room
- Partner in same room, but not same bed
- Partner in same bed

**Questions 48-53: If you have a room mate or bed partner, ask him/her how often in the past month you have had . . .**

48. Loud snoring

- Not during the past month
- Less than once a week
- Once or twice a week
- Three or more times a week

49. Long pauses between breaths while asleep

- Not during the past month
- Less than once a week
- Once or twice a week
- Three or more times a week

50. Legs twitching or jerking while you sleep

- Not during the past month
- Less than once a week
- Once or twice a week
- Three or more times a week

51. Episodes of disorientation or confusion during sleep

- Not during the past month
- Less than once a week
- Once or twice a week
- Three or more times a week

52. Other restlessness while you sleep; please describe:

53. If other restlessness listed in Question 52, then how often has your room mate or bed partner noted this restlessness?

- Not during the past month
- Less than once a week
- Once or twice a week
- Three or more times a week

54. COVID-19 has caused a decrease in my quality of sleep during the past month.

- Strongly agree
- Agree
- Neither agree nor disagree
- Disagree
- Strongly disagree

**Currently...**

55. How many days per week do you exercise?

0 — 7

56. On the days you exercise, how many minutes do you exercise on average?

0 — 360

57. How would you rate your quality of sleep?

0 (worst sleep ever) — 10 (best sleep ever)

58. On average, how many times throughout the week do you feel the inclination to fall asleep during the day?

- Never
- Less than once a week
- Once or twice a week
- Three or more times a week

59. On average, how often do you feel tired or fatigued after your sleep?

- Nearly every day
- 3-4 times a week
- 1-2 times a week
- 1-2 times a month
- Never or nearly never

60. During your waking time, do you feel tired, fatigued, or not up to par?

- Nearly every day
- 3-4 times a week
- 1-2 times a week
- 1-2 times a month
- Never or nearly never

**THANK YOU!**
